# Alterations in Early Alpha-band Connectivity emerge in Infancy among children later diagnosed with Autism

**DOI:** 10.64898/2026.06.25.26353501

**Authors:** Haerin Chung, Winko W. An, Carol L. Wilkinson, Gabriela Davila Mejia, Helen Tager-Flusberg, Charles A. Nelson

## Abstract

Autism is a heterogeneous neurodevelopmental condition, often accompanied by challenges in language and cognitive development. Although atypical functional connectivity (FC) has been reported in autism, the timing of when it first emerges and its relevance for later behavior remain poorly understood. In this study, we examined developmental trajectories of alpha-band FC and network organization across the first three years of life. We computed global alpha-band measures, including peak alpha connectivity frequency (PACF), mean FC, clustering coefficient, and modularity, to characterize nonlinear developmental trajectories from longitudinal EEGs collected from 238 children (3–36-month-olds) with (Autism; n=58) and without (LL-noAutism; n=180) autism. Network-based statistics (NBS-Predict) identified subnetworks contributing to group differences at each age. Exploratory graph analyses (EGA) examined associations among FC, network measures, and language outcomes. We observed that PACF increased linearly with age in both groups. Global alpha-band connectivity measures showed a similar developmental pattern, with mean global FC, clustering coefficient, and modularity all increasing rapidly during the first year in both groups. Thereafter, these measures declined in the Autism group but continued to gradually increase in the LL-noAutism group. Compared to LL-noAutism, NBS-Predict identified both hyper- and hypo-connectivity subnetworks in Autism at 3 months, followed by a hypo-connectivity subnetwork at 24 and 36 months. EGA indicated that early hyperconnectivity predicted later hypoconnectivity and was associated with subsequent network organization and language outcomes. These findings indicate that altered alpha-band connectivity trajectories are detectable in infancy in children later diagnosed with autism and may contribute to later differences in developmental outcomes.

## Introduction

Autism spectrum disorder (ASD) affects approximately 1 in 31 children in the United States^1^. Autistic individuals are highly heterogeneous in their phenotypic presentation of core symptoms (i.e., social communication and restricted repetitive behaviors [RRB]) as well as important developmental milestones such as language development. Many autistic children experience language delays^2^, and approximately 30% remain minimally verbal^3,4^. Early intervention has been found to improve developmental outcomes, yet most autistic children are not diagnosed until approximately four years of age^5^. This delay limits opportunities for intervention during a period of heightened neural plasticity, when the developing brain may be particularly responsive to change. Converging evidence suggests that the neural substrates underlying autism can be detected within the first months of life^6,7^, and early brain differences are thought to contribute to the diverse behavioral profiles observed later, including variation in cognitive function, language development, and sensory processing^8–10^. Deviations in early brain development may exert cascading effects on subsequent brain organization and behavior, underscoring the need to identify when and how these divergences arise.

In typical development, the first six months are marked by rapid maturation, including cortical organization, neuronal specialization, myelination, and dynamic synaptic remodeling^11–13^. Foundational sensorimotor, auditory, and visual networks are established at birth, while higher-order networks such as the default mode and dorsal attention networks mature through the first year^14,15^. These developmental changes can be studied using functional connectivity, defined as the coordinated activity among brain regions, often quantified through statistically dependent measures such as amplitude correlation or phase synchrony^16^. Recent advances in graph theory also provide additional insights into how each network is organized, especially its integration and segregation^17,18^. During early childhood, shifts toward modular architecture (i.e. increased modularity and clustering coefficient reflecting network segregation) allowing efficient information transfer have been documented^19,20^. These changes in large-scale networks are thought to underlie the emergence of increasingly complex cognitive and behavioral skills such as language and executive control^21,22^. The timing of changes in network development can also provide insight on developmental periods vulnerable to atypical network formation^23^.

Growing evidence suggests that individuals with autism exhibit altered functional connections between brain regions that may underlie sensory, cognitive, and social communication challenges. However, *when* these neurodevelopmental processes diverge early in life among children later diagnosed with autism, and *how* such divergence relates to later symptom manifestation is poorly understood. Disrupted excitatory and inhibitory balance has been proposed as a key mechanism^24^, potentially altering sequential stages of neuronal development including synapse formation, circuit assembly, and cortical maturation, leading to atypical connectivity and brain organization^25^. A small number of fMRI studies in early childhood (spanning infancy at 18 months of age to toddlerhood) report atypical functional connectivity and network inefficiencies in autism^26,27^. These include early disruptions in visual and sensorimotor networks^2,28^ in infants with autism, as well as patterns of thalamic-prefrontal underconnectivity and thalamic-occipital and thalamic-motor overconnectivity in from infants at elevated likelihood of developing autism (i.e., those with an older autistic sibling)^2,29,30^. Such early alterations in connectivity potentially contribute to emerging motor, social challenges and repetitive behaviors^31–33^. Yet most neuroimaging work to date has been cross-sectional and focused on older children, and fMRI coactivation patterns lack the temporal precision for assessing the rapid dynamics of neural communication.

EEG coherence offers a complementary, non-invasive, and scalable tool to capture specific changes in brain dynamics and connectivity across infancy and toddlerhood. EEG alpha-band rhythms are considered markers of top-down control and excitatory–inhibitory dynamics^34^, processes that regulate information flow and attentional modulation^35,36^, and have been found to be associated with the structural and functional properties of long-range connections^2,37,38^. However, the functional significance and developmental nature of EEG alpha connectivity in early infancy have yet to be characterized. Across older age groups, widespread reductions in long-range connectivity have been reported in autistic children and adults^9,39–44^. Notably, Dickinson et al.^9^ found decreased frontal but increased temporoparietal alpha connectivity at 3 months, which was associated with autism symptoms at 18 months. Moreover, reports of hyper-alpha connectivity in the first-year contrast with later hypo-alpha connectivity in childhood^45^. These findings are thought to reflect *age-related shifts* that may underlie ongoing processes such as synaptic consolidation^2^, white matter maturation^46^, or thalamocortical development^47^. In addition to connectivity strength, graph theory has also been used to assess connectivity patterns in EEG studies of autism, from which reduced network segregation, including lower clustering coefficients and less optimal information transfer^43,48–50^, has been reported, though exceptions exist^51^.

Despite these advances in autism research using EEG, most work remains cross-sectional and focused on older children or adults, overlooking the dynamic changes in network organization that occur in the first years of life. Research on typical development is similarly limited. Although a few studies^52–54^ have characterized EEG functional connectivity in infants under 24 months, the limited number of overlapping participants constrains the ability to track within-individual developmental change. Consequently, it remains unclear how early EEG connectivity patterns evolve over time within an individual and whether early network differences predict later brain function and behavior. Addressing these questions requires prospective longitudinal studies that track whole-brain connectivity across infancy and toddlerhood.

In addition, most prior EEG studies emphasize global reductions in connectivity, providing limited insight into which specific networks contribute to these differences or how they change over time. Identifying early cortex-wide connectivity patterns that distinguish infants who later develop autism *and* predict later network organization that underlies emerging behavioral function may help clarify the mechanisms through which early brain networks support developmental outcomes. Such markers could also provide clinically meaningful early indicators of autism, supporting earlier diagnosis and intervention aimed at improving developmental trajectories.

### Current Study

This project leverages electroencephalography (EEG) data collected from large-scale longitudinal studies of infants with and without autism to 1) characterize the trajectory of functional connectivity and neural networks in the alpha band in the first 3 years of life, 2) investigate the timing and regional distribution of connectivity differences between young children with autism and their typically developing peers, and 3) explore whether early alterations in connectivity are associated with subsequent differences in connectivity and emerging language skills. Based on previous findings implicating diverging functional connectivity patterns in infancy and toddlerhood in autistic children, we hypothesize that there would be differences in the ***trajectory*** of functional connectivity and network dynamics between autistic children and typically developing peers. Using a within-subject longitudinal dataset, we further explored whether early altered patterns predict later network dynamics as well as individual variations in later emerging language skills.

## Method

### Participants

Participants were recruited as part of the Infant Sibling Project (Study 1, IRB-X06-08-0374) and the Infant Screening Project (Study 2, IRB-P00018377), both prospective, longitudinal studies, enrolling infants with and without first degree family history of autism starting as early as 3-months of age. Specifically, Study 1 recruited infants at either 3 or 6 months of age and followed them longitudinally at 9, 12, 18, 24, and 36 months. Study 2 initially recruited infants at 12–14 months of age, but then added a 3-month enrollment option partway through the study. In addition to infants with a family history of autism, this study also recruited infants with elevated social communication concerns at 12 months of age. All participants were born with minimal gestational age of 36 weeks, birth weight over 2.5kg, and with no known genetic or neurological disorders.

For this study, EEG data were compared between infants later diagnosed with autism (Autism) with low-likelihood infants without autism (LL-noAutism). Our main analysis included 897 EEGs collected from 238 participants (LL-noAutism: *N*=180; Autism: *N*=58; Supplementary Figure 1 and mean age by each age group is reported in Supplementary material Table 1). The LL-noAutism group was restricted to infants without a family history of autism, and infants who did not have elevated social communication concerns at 12 months (from Study 2). The Autism group included all infants later diagnosed with autism either through study assessments at 24 and 36 months of age, or through a community diagnosis. Many infants in the Autism group (n= 49) had an older sibling with autism. Sample characteristics across and within studies are shown in Table 1. Combined, the sample was predominantly white (81.5%), with the majority of mothers having a college education level or above.

**Table 1.** Demographic information and 24-month developmental quotients.

|  | Combined | Autism | LL-noAutism | p-value |
| --- | --- | --- | --- | --- |
| N (%) | 238 | 58 | 180 |  |
| Sex |  |  |  |  |
| Male | 146 (61.3) | 46 (79.3) | 100 (55.6) | 0.002 |
| Female | 92 (38.7) | 12 (20.7) | 80 (44.4) |  |
| Ethnicity |  |  |  |  |
| Hispanic | 11 (4.6) | 7 (12.1) | 4 (2.2) | 0.006 |
| Non-Hispanic | 225 (94.5) | 51 (87.9) | 174 (96.7) |  |
| Not answered | 2 (0.8) | 0 (0.0) | 2 (1.1) |  |
| Race |  |  |  |  |
| White | 194 (81.5) | 47 (81.0) | 147 (81.7) | 0.935 |
| Black | 4 (1.7) | 1 (1.7) | 3 (1.7) |  |
| Asian | 10 (4.2) | 3 (5.2) | 7 (3.9) |  |
| More than one race | 28 (11.8) | 7 (12.1) | 21 (11.7) |  |
| Not answered | 2 (0.8) | 0 (0.0) | 2 (1.1) |  |
| Income |  |  |  |  |
| < \$35,000 | 10 (4.2) | 3 (5.2) | 7 (3.9) | |
| \$35-75,000 | 14 (5.9) | 4 (6.9) | 10 (5.6) | 0.125 |
| > \$75,000 | 191 (80.3) | 41 (70.7) | 150 (83.3) | |
| Not answered | 23 (9.7) | 10 (17.2) | 13 (7.2) |  |
| Education (Maternal) |  |  |  |  |
| ≤ High school/GED | 14 (5.9) | 10 (17.2) | 4 (2.2) | <0.001 |
| Associate Degree | 6 (2.5) | 3 (5.2) | 3 (1.7) |  |
| Bachelor Degree | 59 (24.8) | 18 (31.0) | 41 (22.8) |  |
| > 4 year college degree | 142 (59.7) | 19 (32.8) | 123 (68.3) |  |
| Not answered | 17 (7.1) | 8 (13.8) | 9 (5.0) |  |
| 24-month MSEL Mean (SD) |  |  |  |  |
| Non-verbal | 105.62 (14.41) | 96.70 (13.53) | 108.46 (13.53) | <0.001 |
| Verbal | 110.17 (21.48) | 92.75 (24.18) | 115.75 (17.22) | <0.001 |

### Autism Diagnosis

Autism outcomes were determined using the Autism Diagnostic Observation Schedule^55^ (ADOS-2) and parent–child interaction administered at 24 and/or 36 months of age. During the COVID-19 pandemic, remote autism evaluations for Study 2 included the Brief Observation of Symptoms of Autism^56^ (BOSA), parent–child interaction, and Vineland Adaptive Behavioral Scales-Third Edition, Parent Interview Form^57^ (Vineland-3). Notably, the convergence between BOSA and ADOS-2 has been established^58^. For toddlers meeting criteria on the ADOS-2/BOSA or coming within three points of cutoffs, a licensed clinical psychologist or developmental behavioral pediatrician reviewed assessment scores and videos and provided their best clinical judgment regarding an Autism diagnosis using the DSM-5 criteria.

### Developmental Assessment

The Mullen Scales of Early Learning^59^ (MSEL) is a standardized developmental assessment for children 0-68 months of age. The verbal developmental quotients were calculated based on the participant’s average age equivalents across subscales (non-verbal: Fine Motor, Visual Reception; verbal: Expressive Language, Receptive Language) divided by the child’s chronological age multiplied by 100.

### EEG Collection, Preprocessing, and Analysis

#### EEG Data Acquisition

For both studies, 2-5 minutes of resting-state baseline EEG were acquired while infants sat on a caregiver’s lap in a dimly lit, sound-attenuated room. Infants either watched a research assistant blow bubbles or a silent video of abstract moving shapes (see SI1 for more details on data collection). Study 1 used 64- or 128-channel Geodesic Sensor Nets (Electrical Geodesic Inc.) connected to a NetAmps 200 or 300 amplifier, sampled at 250 or 500 Hz. Study 2 used only 128-channel nets with a NetAmps 300 amplifier at 500 Hz. EEG was collected with a 0.1 Hz high-pass analog filter, online re-referenced to Cz, with impedances below 100 KΩ. No significant differences were observed between NetAmps 200 and 300 collection parameters^10^.

#### EEG pre-processing

EEG data were first spatially downsampled to a subset of channels spanning the entire scalp to harmonize recordings across net types (see Supplemental Figure 2). Data preprocessing and data quality assessment was carried out via the Harvard Automated Processing Pipeline for EEG^60,61^ (HAPPE v3.0). Each EEG was bandpass filtered (1-100 Hz) and resampled to 250 Hz. Electrical line noise was removed at 60 Hz via CleanLine. Artifacts were rejected (e.g., eye blinks, movement) via wavelet-thresholding. The EEG data were then re-referenced to the average reference and segmented into contiguous 1-s windows with 50% overlap. Segments with signal amplitude exceeding the −150 to +150 μV range were additionally removed.

#### EEG rejection criteria

EEG recordings were rejected if they contain <60 segments^62^ or <80% percent good channels. These quality metrics were not significantly different across outcome groups (Supplemental Table 2). Of 956 EEG recordings that were collected, 21 did not meet these criteria and were excluded. Participants with usable data from only a single time point were excluded from the trajectory analyses, as at least two observations are required to estimate changes within an individual.

#### Functional Connectivity Analyses

We first transformed the preprocessed EEG signals using current source density (CSD) with MNE-Python^63^ to mitigate confounding effects of volume conduction and improve accuracy of functional connectivity analyses. CSD applies a surface Laplacian, which computes the second spatial derivative of the scalp potential across electrodes, reducing the spatial spread of neural sources. Coherence (COH) was then calculated between each pair of electrodes (e.g., signal x and y) at each frequency f using MNE-Connectivity^64^ with the following formula:

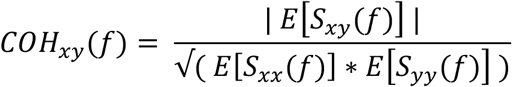

where S_xy_(f) is the cross-spectral density between *x* and *y*, and S_xx_(f) and S_yy_(f) are power spectral densities of x and y, respectively. All power spectra were estimated using a multitaper method with a time-bandwidth product of 1 (mt_bandwidth=1). Essentially, COH reflects the consistency of phase differences between signals *x* and *y* across all trials, ranging from 0 (no consistency) to 1 (perfect consistency). We acknowledge that certain measures, such as those from the phase lag index (PLI) family, reduce the influence of zero-lag connectivity to minimize the effects of volume conduction. However, given that zero-lag connectivity may represent both artificial (volume conduction) and true (synchronization between neural signals) connectivity, excluding zero-lag phase connectivity represents a sensitivity-specificity trade-off: reducing spurious connectivity at the potential cost of discarding physiologically meaningful connectivity. Here, we chose COH to maximize sensitivity^65^, and applied CSD transformation prior to COH calculation to improve specificity by reducing spatial leakage and volume conduction between neighboring sensors. For each participant, a total of 1,275 COH values—representing all possible pairs among 51 electrodes—were computed at each frequency point between 1 and 40 Hz, with a resolution of 0.2 Hz. Each set of the 1,275 values was organized into a COH matrix, symmetric across its main diagonal (see Figure 1A, B).

**Figure 1.**
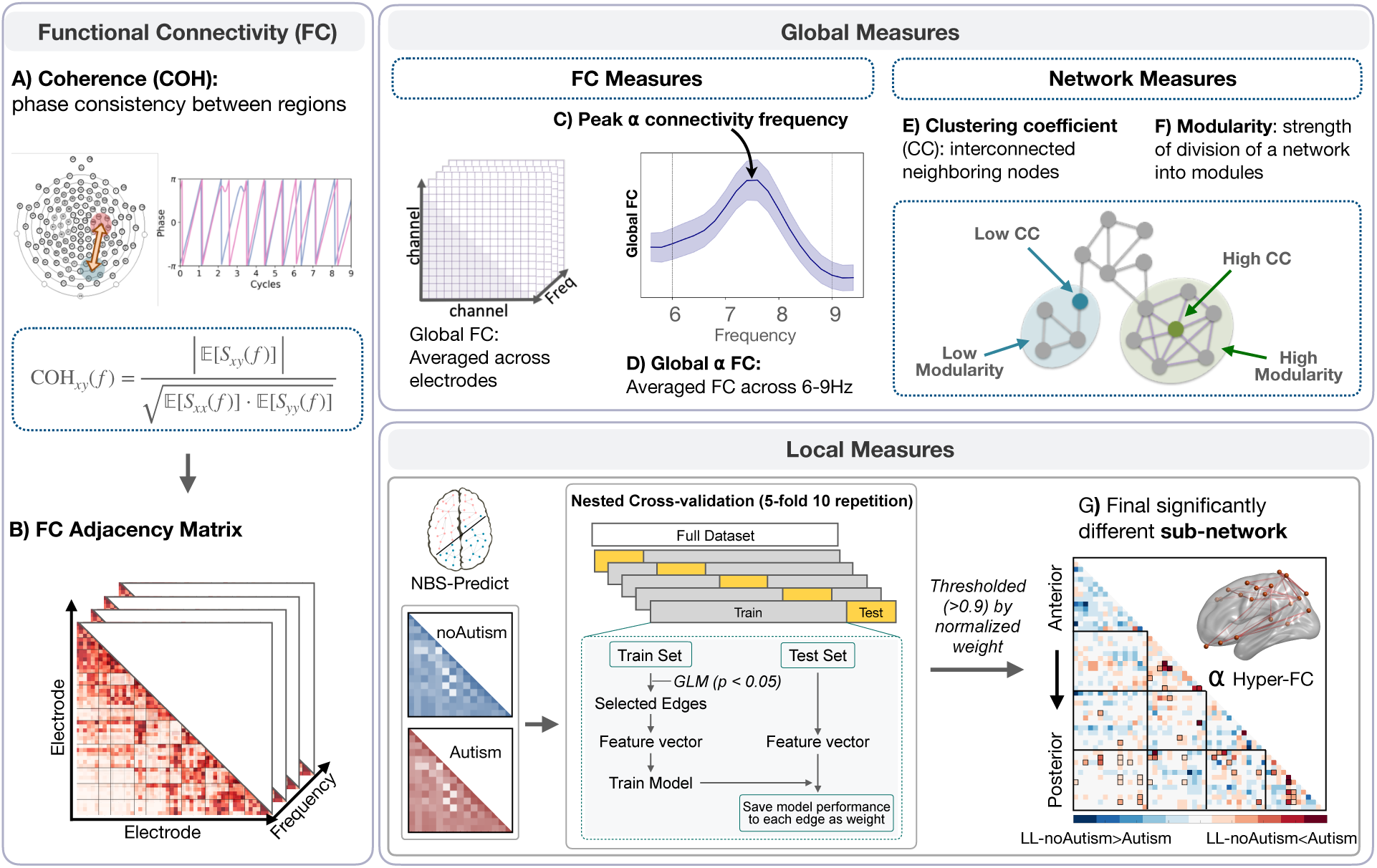
Overview of methods. Panel A) Functional connectivity (FC) was estimated using coherence (COH), a measure of consistency in phase differences between electrode pairs, defined as the normalized cross-spectral density between signals. Panel B) Pairwise COH values were organized into FC adjacency matrices for each participant per frequency. Global FC measures were extracted in the alpha band (Panels C-F), including peak alpha connectivity frequency (Panel C) and mean global alpha FC (Panel D). Graph-theoretic network measures were computed, comprising the clustering coefficient (CC), reflecting the degree of local interconnectedness among neighboring nodes (Panel E), and modularity, quantifying the extent to which the network segregates into distinct functional modules (Panel F). To identify locally significant connectivity differences between groups, a nested cross-validation framework (5-fold, 10 repetitions) was implemented using NBS-Predict. Within each training fold, a general linear model selected edges that significantly differentiated groups, which were used as features to train a classifier. Edge-wise model performance from each iteration was saved as a weight and normalized across all iterations. Connected edges with a normalized weight surpassing a threshold of 0.9 were retained, yielding a final sub-network of significantly different connections (Panel G) for each age group.

#### Global measures

##### Global COH spectrum and Peak alpha connectivity frequency

For each participant, all COH values were averaged across electrode pairs at each frequency point to derive the global COH spectrum. The peak alpha connectivity frequency (PACF) was defined as the frequency within the 6–10 Hz range at which the global COH reached its highest peak (Figure 1C), identified using the find_peaks function from SciPy. We assessed PACF starting at 170 days and beyond, as the prior literature indicates that most infants start to exhibit a single robust dominant peak at this age^47,66^ (See supplemental material Table 3 for the full range and median of PACF by age group.) The magnitude of mean alpha functional connectivity (Figure 1D) was computed as the average across all possible pairs in the lower diagonal of connectivity matrices in the alpha band (6–9Hz, as the median of PACF were between 7.4 and 8.4Hz, and to be consistent with prior literature on infant-toddler EEG alpha-band coherence)^66–68^.

##### Graph theoretical analysis in alpha band

The alpha-band COH matrix for each participant was obtained by averaging individual COH matrices across the 6–9 Hz frequency range. To analyze the network structure, we first binarized each matrix by applying a threshold set at the 70th percentile of its non-diagonal values. From the resulting binary adjacency matrix, we computed key graph-theoretical metrics using the NetworkX Python package^69^, including clustering coefficient (i.e. quantifying the degree to which nodes in the network tend to form locally interconnected clusters), and modularity (i.e. strength of the division of the network into these communities, with higher values indicating more pronounced community structure; see Figure 1E, F).

#### Local measures

To identify specific edge-level differences in groups, a series of edge analysis were conducted using Network-Based Statistic-Predict^70^ (*NBS-Predict*). NBS-Predict is a prediction-based extension of the NBS framework designed to identify brain network connections that are predictive of behavioral or clinical outcomes. It integrates machine learning with cross-validation to directly assess the predictive utility of connectivity patterns at the network level, and permutation testing is employed to assess the statistical significance of predictive performance while controlling for multiple comparisons across the network.

We employed NBS-Predict for Autism vs LL-noAutism classification at each age separately. We selected Logistic Regression as the classifier and set the following parameters: k-fold=5, repetition=10, permutation=500, scaling method=StandardScaler, metric=F1, edge threshold=0.9. For each age, we ran this method twice, once for the contrast of “Autism>LL-noAutism”, and the other for “LL-noAutism>Autism”, and then deemed a model having good predictive power if its p<0.05. This procedure identified four significant clusters: both “Autism>LL-noAutism” and “LL-noAutism>Autism” at 3 months, and “LL-noAutism>Autism” at 24 and 36 months (see Figure 1G; Table 2). These clusters were subsequently used as “statistical masks”, within which functional connectivity values were averaged to derive local measures representing age-specific group differences.

**Table 2.** NBS-Predict model performance in ASD vs TD classification at each age.

| Age | Contrast | F1 | p | Nodes | Edges | NBS subnetwork |
| --- | --- | --- | --- | --- | --- | --- |
| 3 | **LL-noAutism > Autism** | **0.55** | **0.004** | 39 | 102 | **Early-hypoconnectivity** |
|  | **Autism > LL-noAutism** | **0.51** | **0.020** | 27 | 80 | **Early-hyperconnectivity** |
| 6 | LL-noAutism > Autism | 0.33 | 0.444 | 5 | 8 | - |
|  | Autism > LL-noAutism | 0.26 | 0.778 | 1 | 0 | - |
| 9 | LL-noAutism > Autism | 0.31 | 0.548 | 3 | 4 | - |
|  | Autism > LL-noAutism | 0.41 | 0.252 | 5 | 10 | - |
| 12 | LL-noAutism > Autism | 0.34 | 0.210 | 8 | 14 | - |
|  | Autism > LL-noAutism | 0.34 | 0.074 | 15 | 30 | - |
| 18 | LL-noAutism > Autism | 0.36 | 0.120 | 8 | 18 | - |
|  | Autism > LL-noAutism | 0.33 | 0.146 | 3 | 4 | - |
| 24 | **LL-noAutism > Autism** | **0.44** | **0.032** | 35 | 94 | **Later-hypoconnectivity** |
|  | Autism > LL-noAutism | 0.40 | 0.056 | 4 | 6 | - |
| 36 | **LL-noAutism > Autism** | **0.48** | **0.014** | 47 | 242 | **Later-hypoconnectivity** |
|  | Autism > LL-noAutism | 0.35 | 0.552 | 3 | 4 | - |

### Statistical analysis

Using global and local measures derived from children with and without autism, we examined group differences in their developmental trajectories over time.

#### Main Analysis: Generalized additive mixed models (GAMMs)

Developmental trajectories were modeled using Generalized Additive Mixed Models (GAMMs; mgcv v1.9-1, R v4.3.2), which accommodate nonlinear changes and repeated measures. GAMMs have proven useful for exploring relationships between measures across time, particularly where the patterns are not yet well established (e.g. between the measure of interest and age^47,71^ as well as unbalanced longitudinal data^72^). Models included outcome group, a smoothed age term (k=4), study and participant as random effects, and sex as a covariate. An age-by-outcome interaction term was retained when it significantly improved model fit (p < .05). To correct for multiple comparisons, the false discovery rate (FDR) was controlled using the Benjamini and Hochberg method, which was applied for each model term to produce q-values (FDR-corrected p-values; see SI2 for more details on GAMM models).

To further understand the nonlinear trajectories of change, inflection points were calculated using the argrelextrema function from scipy in python with order=100. A standardized rate of change per day was calculated to visualize developmental changes within features. The modeled value of a feature at a given age (in days) was subtracted from the modeled value from the subsequent day, and this was divided by the standard deviation of the modeled values of that feature across the age range.

#### Exploratory Analysis: Exploratory graph analysis (EGA)

As an exploratory analysis, we examined whether early connectivity differences contributed to group-level differences observed later in toddlerhood and whether these early differences were associated with subsequent developmental skills. EGA analysis was used to characterize variables and their unique relationships (associations)^73,74^. A key strength of EGA lies in its ability to detect the existence and weight of edges, which is computed with the conditional associations between variables—statistical relationships between two variables that remain after taking accounting for all other variables in the data (e.g., partial correlations). EGA was carried out applying the graphical least absolute shrinkage and selection operator (GLASSO) and estimated using a penalized maximum likelihood solution based on the extended Bayesian information criterion. We conducted EGA with a subset of individuals (N=30; LL-noAutism: 18, Autism: 12) with EEG data at ***both*** 3 and 24 months ***and*** with MSEL verbal DQ at 24 months. We limited the analysis to 3 and 24 months because the data collection at 24-36 months was substantially disrupted by the COVID-19 pandemic for Study2. Accordingly, given the small sample, we acknowledge that the findings of this analysis should be interpreted with caution.

## Results

Longitudinal resting state EEG was analyzed from infants with and without a later diagnosis of autism. Participants were aged 3-40 months, across 2 studies conducted in the same laboratory. Notable changes in functional connectivity were observed across age bins and between outcome groups. To further characterize these developmental changes and to assess whether the trajectory differed between outcome groups, we used generalized additive mixed models (see Supplemental Figures 3,4 for Figures based on non-GAMMs modeled measures). In total, we conducted four independent GAMMs to predict each global measure (PACF, mean alpha functional connectivity, clustering coefficient, and modularity).

### Global measures: Functional connectivity and network characteristics

GAMMs revealed significant age-dependent changes across all four global measures, with divergent developmental trajectories between the Autism and LL-noAutism groups for mean alpha functional connectivity, clustering coefficient, and modularity (Figure 2; Supplementary Table 4). PACF increased linearly across the first three years of life in both groups, with no significant group differences in trajectory (F = 56.65, q < 0.001; age-by-outcome interaction p > 0.2)). In contrast, mean alpha functional connectivity, clustering coefficient, and modularity showed significant age-by-outcome interactions (q <0.01 for all).

**Figure 2.**
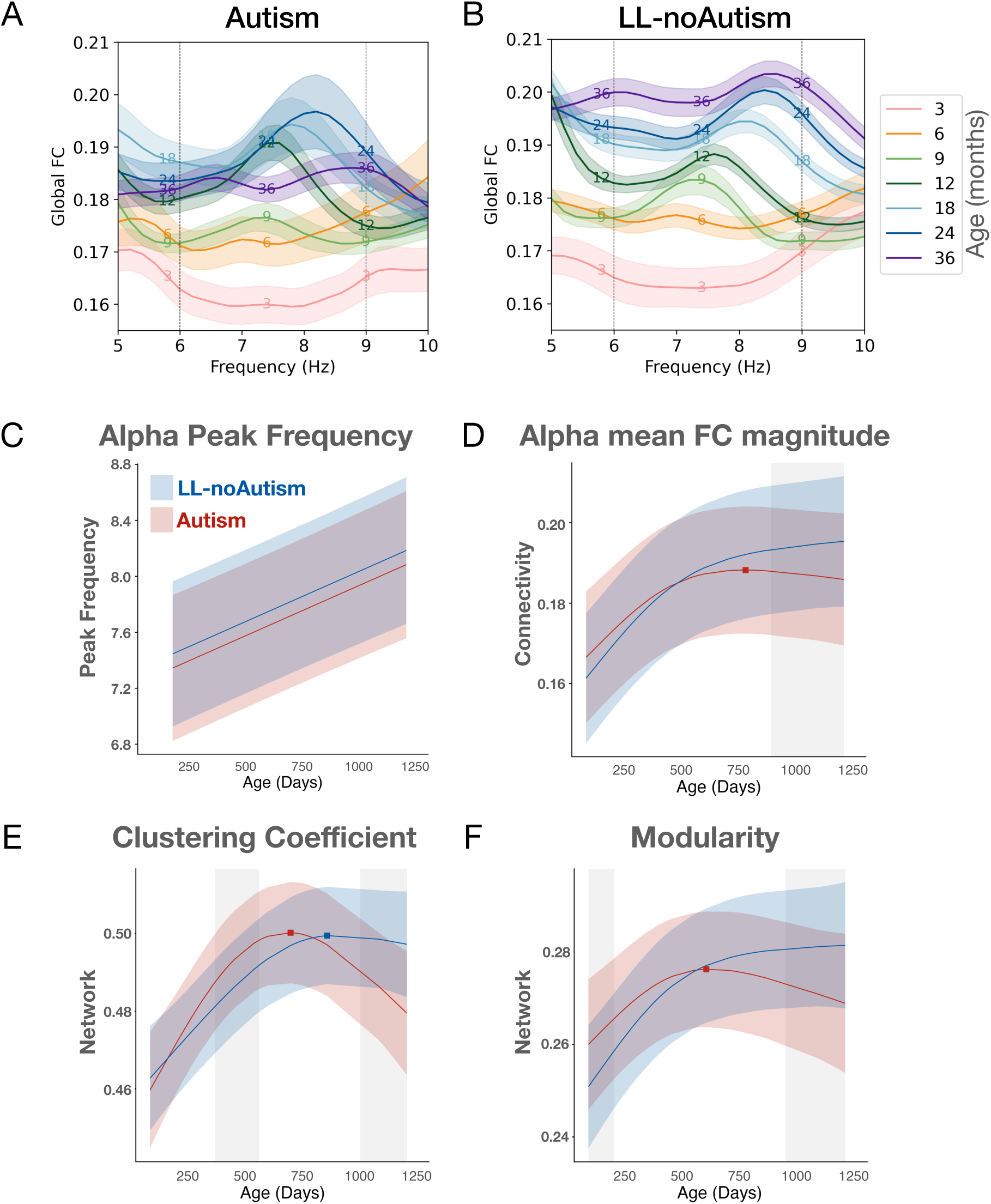
Panel A) Global functional connectivity (averaged across all possible pairs) spectra visualized across theta and alpha frequency ranges across 7 age bins. Global functional connectivity from EEGs collected within each age bin were averaged and shading denotes 95% confidence intervals. A: LL-noAutism; B: Autism. Panels C-F: GAMMs modeled trajectories for LL-noAutism (blue) and Autism (red) for Panel C) PACF, Panel D) mean functional connectivity, Panel E) clustering coefficient, and Panel F) modularity. Lines are the model predicted value with the shaded area representing 95% confidence intervals. Relative inflection points are shown with circular (min) and square (max) markers.

For alpha functional connectivity, both groups showed comparable linear increases through approximately 26 months, after which trajectories diverged: the Autism group exhibited a decline, while the LL-noAutism group continued to increase gradually through 36 months. Clustering coefficient and modularity showed complementary patterns, with the Autism group reaching earlier peaks followed by pronounced declines, whereas the LL-noAutism group sustained more stable trajectories into later childhood. Notably, modularity and clustering coefficient was transiently higher in the Autism group early in infancy that declined after age 2, suggesting reduced network segregation.

### Local measures: Hyper- and hypo-connectivity subnetworks in Autism

We conducted NBS-Predict analyses at each age visit to detect subnetworks composed of edges that contributed to predicting later autism diagnosis. NBS-Predict identified four significant clusters (subnetworks): both “Autism > LL-noAutism” and “LL-noAutism > Autism” at 3 months, and “LL-noAutism > Autism” at 24 and 36 months (Table 2). For clarity, we refer to these as the ***early-hyper*** (Autism > LL-noAutism), ***early-hypo*** (LL-noAutism > Autism), and ***later-hypo*** (LL-noAutism > Autism) NBS subnetwork connectivity measures at 24 and 36 months, with reference to the Autism group (Figure 3A, B). Total node degree (i.e., number of total connections) for each electrode by age are presented in Supplemental Tables 5-7.

**Figure 3.**
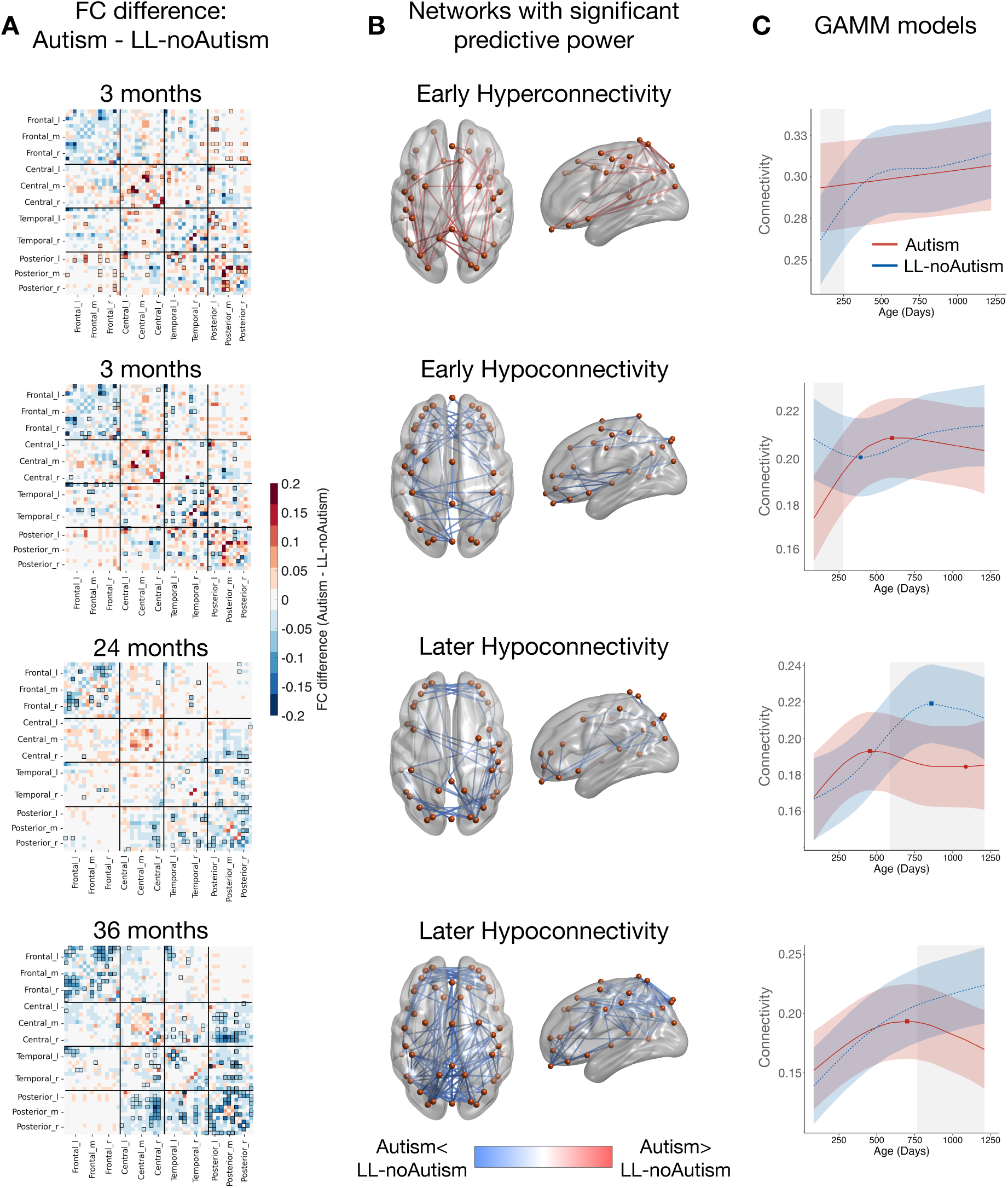
Panel A) Adjacency matrix showing the NBS-defined significantly different pairs that between groups. Panel B) Visualization of the significantly different NBS-defined subnetworks between groups (weighted threshold = 0.9) on a 3D brain surface generated by the BrainNet Viewer. Panel C) GAMMs modeled trajectories for LL-noAutism (blue) and Autism (red) for NBS-defined subnetworks. COH values were averaged across all off-diagonal entries within the corresponding black mask areas in section A for this analysis. Lines are the modeled mean predicted value with the shaded area representing 95% confidence intervals. Relative inflection points are shown with circular (min) and square (max) markers.

At 3 months, we discovered two significant subnetworks. First, ***early-hyperconnectivity*** suggested that infants later diagnosed with autism exhibited significantly stronger connectivity primarily in posterior-to-posterior, frontal-to-posterior, and central-to-central connections (F1= 0.51, adj-p = 0.02). These patterns of posterior over-connectivity are consistent with findings from the GAMMs analysis of region-to-region FC, where increased connectivity in infancy was observed in central-to-posterior and posterior-to-posterior pairs and (see SI3-Local features: functional connectivity by ROI and Supplemental Figure 5). In contrast, ***early-hypoconnectivity*** is characterized by reduced connectivity in the Autism group involving frontal-to-frontal and frontal-to-temporal connections (F1= 0.55, adj-p = 0.004).

Across 24 and 36 months, ***later-hypoconnectivity*** comprised of a persistent pattern of reduced frontal and posterior connectivity in autistic toddlers. This pattern emerged at 24 months (F1= 0.44, adj-p = 0.032) and by 36 months, this under-connectivity in the Autism group became more diffuse (Figure 3A,B). That is, a larger subnetwork (a greater number of identified nodes and edges) was identified as exhibiting significantly lower FC relative to the LL-noAutism group (F1= 0.48, adj-p = 0.014), encompassing frontal-to-frontal connections and extended reduced connectivity across posterior, temporal, and central regions. Again, these patterns of frontal and posterior hypo-connectivity in Autism are consistent with findings from the GAMM analysis of local features (Supplementary Figure 5). Visual inspection of the FC difference adjacency matrix indicated an increasingly reduced frontal connectivity, particularly across left-right regions, suggesting reduced interhemispheric frontal connectivity in the Autism group relative to LL-noAutism group across the first three years (Figure 4).

**Figure 4.**
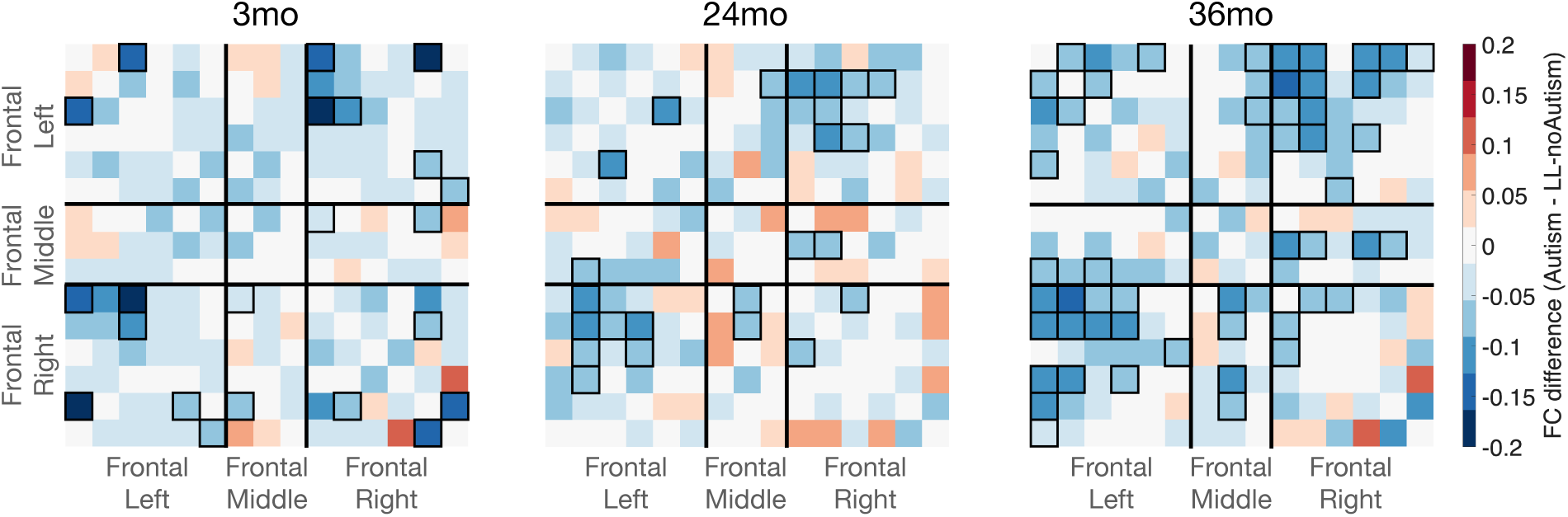
Adjacency matrix showing hypoconnectivity within the frontal region in autism at 3, 24, and 36 months. Significant differences between the Autism and LL-noAutism groups (NBS-defined) are highlighted by black squares. More significant edges correspond to interhemispheric connections.

#### Trajectory of NBS identified clusters

First, the trajectory of **early hyperconnectivity at 3 months** exhibited a significant age-by-outcome interaction (Supplemental Table 4; F = 17.68, q < 0.001). Autistic infants showed elevated connectivity during early infancy that persisted until approximately 8 months, with connectivity increasing modestly with age (Figure 3C). Second, the subnetwork representing **early-hypoconnectivity at 3m** revealed a diverging pattern between Autism and LL-noAutism group, where autistic infants exhibited hypoconnectivity up until 9 months. The autism group exhibited a gradual increase during the first year, peaking in the second year followed by a decline. In contrast, the LL-noAutism group showed less changes across age, with a slight decrease followed by increase in the second year. Third, the trajectory of **later-hypoconnectivity at 24 and 36 months** showed patterns consistent with global measures. The LL-noAutism group showed a gradual increase during the first year, peaking in the second year. By contrast, the Autism group showed a ‘growth-then-decline’ trend (Figure 3C).

### Exploratory analyses: Association between NBS identified subnetwork connectivity and behavior

First, EGA indicated a negative association between NBS early-hyperconnectivity subnetwork at 3 months and NBS later-hypoconnectivity subnetwork at 24 months (Figure 5A,B). Infants with stronger early-hyperconnectivity tended to exhibit weaker connectivity in the later-hypoconnectivity subnetwork during the second year. Second, later-hypo NBS subnetwork connectivity was also positively associated with modularity at 24 months. Finally, although these analyses were exploratory given the modest sample size, these features tracked variability in language skills, such that higher early-hypo NBS subnetwork connectivity at 3 months was associated with lower verbal developmental quotient (Figure 5C). Together, these patterns suggest that early differences in functional connectivity in infants later diagnosed with autism may influence the subsequent developmental organization of large-scale networks, including networks relevant for emerging complex skills such as language.

**Figure 5.**
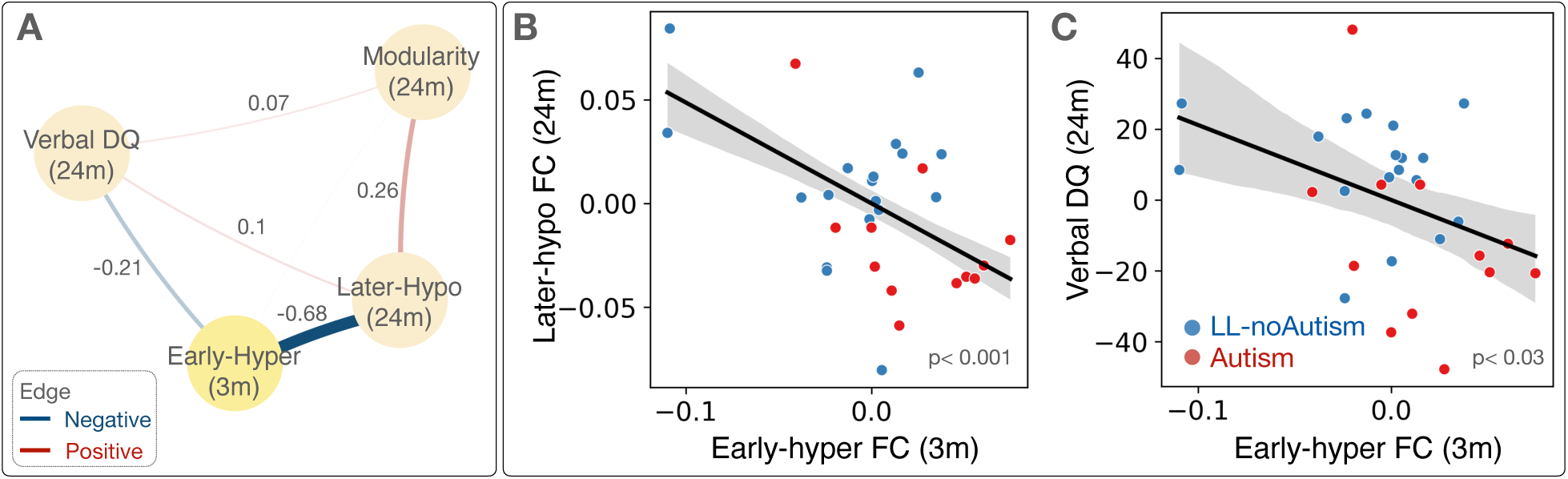
Panel A) Network structure from EGA (n = 30). Circles indicate variables, and lines indicate association between variables (blue: negative association, red: positive association). Thicker lines indicate greater magnitude of the association (Edge-weights are reported alongside the edges). Scatterplots representing association between Early-hyperconnectivity at 3m and Later-Hypoconnectivity at 24m (Panel B) and Verbal DQ at 24m (Panel C). (*Note.* The associations in (B) and (C) reflect residualized associations by study and sex.)

## Discussion

Leveraging two longitudinal datasets with a prospective study design, we characterized the emergent trajectory of EEG alpha-band functional connectivity and network organization during infancy and toddlerhood. Our results revealed distinctive patterns between typically developing infants and infants diagnosed with autism, such that infants with autism exhibited reduced global alpha connectivity during the second and third years, complemented by earlier peaks and subsequent reductions in efficient network organization (i.e., clustering coefficient and modularity). Edge-level analyses revealed central-posterior hyperconnectivity and frontal hypoconnectivity at 3 months, followed by reduced interhemispheric frontal connectivity in the autism group throughout infancy to toddlerhood. Finally, we found that early hyperconnectivity was associated with later hypoconnectivity and lower language scores in toddlerhood across groups, suggesting that early alterations in connectivity may have lasting impacts on network development and behavior.

## 1: Trajectory of global alpha measures in typically developing (TD) children

TD children showed increases in global alpha-band connectivity measures (i.e., peak alpha frequency, global functional connectivity, clustering coefficient, and modularity), supporting improved network segregation over the first years of life. These increases were steeper during the first year than in the second or third years. Although spectral EEG measures do not directly index structural maturation, this trajectory is temporally aligned with the development of important brain structures and networks. We therefore propose a hypothesis regarding the underlying processes linked with the observed changes in alpha-band functional connectivity.

The developmental trajectory of EEG alpha-band functional connectivity observed in our study closely parallels known patterns of grey and white matter maturation in early childhood^75–77^, both of which are fundamental for establishing and strengthening thalamocortical circuitry. During the first years of life, the thalamocortical system undergoes rapid expansion, from early synaptic proliferation, increased neuropil volume, and progressive myelination to a gradual reorganization and slowing down in growth^75,78^. These structural changes may shape alpha activity as there is converging evidence linking alpha oscillatory properties to both grey and white matter development. For example, the amount of neuropil and active synapses has been positively associated with alpha band power^2^. Increased axonal myelination and improved tract organization, supporting more efficient thalamocortical communication^79^, have also been associated with increases in peak alpha frequency^2,80^. Conversely, weaker or atypical white matter tracts, such as in autism^81^, may reduce the stability or synchrony of long-range alpha connectivity and diminish cross-regional coordination^82^. Taken together, we hypothesize that the observed increases in alpha functional connectivity across infancy and toddlerhood reflect, at least in part, the underlying maturation of thalamocortical pathways as shaped by coordinated grey and white matter growth.

## 2. Altered alpha-band connectivity in autism

Distinct EEG trajectories were observed in infants later diagnosed with autism, characterized by an early phase of hyperconnectivity followed by a later phase of hypoconnectivity among sensorimotor regions (posterior and central). In contrast, in frontal regions, consistent hypoconnectivity was observed, driven by reduced interhemispheric communication. These findings suggest atypical maturation of large-scale networks, with excessive early “cross talk” between sensorimotor networks as well as reduced long-range frontal communication, which may causally disrupt subsequent network specialization and brain development^83,84^.

### 2.1 Early hyperconnectivity and Later hypoconnectivity among sensorimotor regions

Edge-level analyses at 3 months revealed hyperconnectivity in infants later diagnosed with autism between visual and sensorimotor regions, a pattern that parallels prior MRI findings in autistic children^2,85,86^ and in autistic siblings^87^. Increased coupling among sensory systems early in development has been reported, and this heightened early “cross-talk” may interfere with the specialization of these systems, each of which undergoes development within distinct sensitive periods characterized by heightened plasticity (Damme et al., 2025). The onset and closure of sensitive periods in sensory systems are regulated by excitatory–inhibitory balance^84,88–90^, which also coordinates synchrony across distributed neural populations through alpha-band oscillations^34,91^. Alpha band rhythms are mediated in part on thalamocortical circuits that support integration of sensory information^92^. As autism has been hypothesized to involve disruptions or delays in inhibitory circuit maturation, the observed alpha hyperconnectivity may reflect alterations in excitatory–inhibitory balance and associated thalamocortical network dynamics. Such alterations could potentially disrupt the timing of sensitive periods and limit gradual network development^93–95^.

Consistent with this interpretation, increased functional connectivity between visual and sensorimotor networks in toddlers with ASD has been associated with symptom severity^28^, and atypical multisensory processing has been linked to downstream differences in social and communicative behaviors^96^. Altered trajectory in somatosensory regions has further been suggested to underlie higher-order language processing^87^. Early hyper- and later hypoconnectivity in our data appear to mirror the documentation of disrupted timing and differentiation of functional networks in autism^28,97^, which could also constrain specialization in networks, and thus, have long-term impacts on later development of brain networks.

### 2.2 Interhemispheric hypoconnectivity within the frontal region throughout the first 3 years of life

In contrast to early central-posterior hyperconnectivity, infants later diagnosed with autism showed persistent interhemispheric hypoconnectivity within frontal regions across the first three years of life. In our prior work, we had reported greater developmental change in frontal aperiodic activity (i.e., an indirect measurement of E/I balance) during the first year of life in infant siblings later diagnosed with autism^10^. These findings align with theoretical models of developmental perspectives in autism^83,98^, proposing that early disruptions to GABAergic and glutamatergic systems may heighten neural noise and alter activity dependent network maturation and specialization^24^, then impacting premature development of frontal networks^99^. Within this framework, we speculate that early posterior hyperconnectivity may be a correlate for atypical early-stage sensory processing, whereas reduced frontal connectivity may indicate altered development of later-emerging anterior systems that modulate and refine large-scale networks. In particular, the prefrontal cortex develops over a prolonged period and exerts top-down influence on sensorimotor systems^98^. Reduced interhemispheric frontal connectivity observed in our study may contribute to diminished anterior modulation, and thus, associated with the observed shifts from early hyper- to later hypoconnectivity. Our findings align well with previous studies that have reported early hyper-sensorimotor connectivity to hypo-prefrontal connectivity infants with heightened likelihood of autism^100^, as well as the studies that have documented altered white matter tracts in frontal regions in infants who have autistic siblings as early as 1.5 months of age^76^.

### 2.3 Developmental cascade framework

Together, our results indicate that group differences are not static but emerge at specific developmental time points and support a developmental cascade in autism: early overconnectivity may reflect premature integration across sensorimotor and visual cortical systems, which in turn hinders the formation and fine-tuning of efficient, modular, specialized networks. <u>Crucially, we hypothesize that it may be the ***timing*** of</u> <u>these early disruptions (when alterations are observed), rather than the presence of</u> <u>differences alone, that drives downstream alterations in cognitive development and</u> <u>behavior</u>. Alpha-band dynamics in particular may offer a sensitive early marker of disrupted large-scale network maturation in autism, capturing deviations that precede and potentially shape both later neural and behavioral outcomes.

Interpreting connectivity within this developmental framework also helps reconcile previously inconsistent findings. Prior studies have reported both hyper- and hypoconnectivity in autism, often interpreted as conflicting results (see O’reilly et al., 2017^101^ for review). Our data suggest that these patterns may instead indicate transient, developmentally specific changes across regions and age. Our results further highlight the value of collecting and analyzing prospective longitudinal data for identifying when divergence occurs and how early differences cascade into later network reorganization. Future studies would benefit from using normative developmental models of functional networks that characterize individual deviations from age-expected trajectories. Such models would allow individual-level precision for early identification and screening of atypical maturation, as well as personalized post-intervention monitoring^102^.

## 3. Implications for Early Detection

With NBS-Predict, we demonstrated the potential utility of identifying neuroimaging-based early biomarkers, revealing a subnetwork of connected edges that distinguished infants later diagnosed with autism at 3m, a period that far precedes behavioral manifestations. Despite the modest sample size, NBS-Predict enabled out-of-sample evaluation of suprathreshold components for individual-level prediction applying a machine learning approach. EEG alpha-band connectivity offers a mechanistically grounded, scalable approach for tracking network development in early life, capturing both global and local patterns of activity that align with fMRI-derived functional resting-state networks and core neurobiological processes such as E/I dynamics and thalamocortical circuit maturation. Our findings indicate that alterations in cortico-cortical organization are detectable as early as 3 months in infants later diagnosed with autism, in line with converging evidence from power-based analyses^7,103^. Notably, the subnetwork edges identified via NBS-Predict were specifically associated with later language development. Together, these findings support the feasibility of identifying neural biomarkers of autism before the emergence of overt behavioral symptoms. Detecting these early differences may improve earlier stratification and support earlier interventions during a period of heightened neuroplasticity.

## 4. Limitations

This study has several limitations. First, sample sizes were reduced at later time points due to the COVID-19 pandemic, affecting the 24- and 36-months assessments in the Study 2 sample. Although our findings reveal interesting developmental patterns, they require replication in a larger longitudinal sample. Second, the findings are limited in spatial interpretation. EEG-derived functional connectivity provides indirect measures of large-scale neural coordination but does not allow precise localization of underlying sub-cortical generators. Studies using source localization approaches or multimodal imaging, such as simultaneous fNIRS, could help improve spatial specificity. Third, the estimation of functional connectivity in this study is defined as statistical dependencies between signals and does not imply directed or causal interactions. Future work should examine the developmental trajectory of effective connectivity to better characterize directional influences within emerging brain networks and to support casual network (re)organization. Finally, our current study is limited to identifying correlational associations between early-emerging brain differences and later autism outcomes and therefore cannot establish causal mechanisms underlying autism. Future work should investigate the specificity of these findings, whether the observed patterns are unique to autism and associated language delays or whether they reflect broader neurodevelopmental processes that overlap with other conditions.

## Conclusion

Early differences in the trajectory of alpha-band functional connectivity were evident in toddlers with and without autism. Connectivity patterns in infancy were associated with connectivity patterns observed later in toddlerhood and language skills, suggesting that early alterations in connectivity observed in autistic toddlers may influence subsequent impact on developmental outcomes, including language ability.

## Author Contribution

H.C., W.W.A., and C.L.W. designed research; H.C., W.W.A., H.T.-F., and C.A.N. acquired funding and resources; C.L.W., H.T.-F., and C.A.N. supervised the project; H.T.-F., and C.A.N. performed investigation and performed project administration; H.C., W.W.A., C.L.W., and G.DM. curated data; H.C., W.W.A., conceptualized the project, performed methodology, created the software, performed formal analysis and visualization; G.DM., C.L.W., H.T.-F., and C.A.N. reviewed and edited the paper; and H.C. and W.W.A. wrote the paper.

## Acknowledgements

We thank all the children and families who participated in this research. We thank all the research staff involved in participant recruitment, data collection, and database administration. This research was supported by the National Institutes of Health (R01-DC010290 to C.A.N. and H.T.F.), the Rosamund Stone Zander and Hansjoerg Wyss Translational Neuroscience Center at Boston Children’s Hospital (to W.W.A.), and Thrasher Early Career Award (to H.C.).

## Data Availability

The data that support the findings of this study are available on request from the corresponding author. The data are not publicly available due to privacy or ethical restrictions.

## Code Availability

The code used to compute EEG functional connectivity matrices and for statistical analyses can be found: https://osf.io/wc2ud/?view_only=b30e552ea6c849d097c64655a9a5df34 (anonymized for peer review).

