## Supplementary Material for "Alterations in Early Alpha-band Connectivity emerge in Infancy among children later diagnosed with Autism"

*Affiliations where work was completed*

1 Division of Developmental Medicine, Boston Children's Hospital, Boston, MA, USA.
2 Harvard Medical School, Boston, MA, USA.
3 Department of Psychological and Brain Sciences, Boston University, Boston, MA, USA.
4 Harvard Graduate School of Education, Cambridge, MA, USA.
*Designates co-first authors

^Corresponding author:
Charles A. Nelson, PhD
Address: BC524, 2 Brookline Place, Brookline, MA USA 02445


### Extended methods

Here we provide supplemental information (SI) to accompany the main manuscript. SI1 provides details on our EEG data processing and analysis. SI2 we discuss additional information on statistical analyses. SI3 we included results from additional analyses for local edge-level findings.

### SI1. EEG data collection and processing procedures

#### EEG Data Acquisition

In Study 1, a research assistant kept infants calm by blowing bubbles and/or showing toys during recording. In Study 2, infants watched a silent video of abstract moving shapes; research assistants avoided social interaction but occasionally blew bubbles across the room or offered a quiet toy if the infant became fussy. Study 1 included recordings with either a 64-channel Geodesic Sensor Net (n = 52; < 6% of the current dataset; 22% of Study 1) or a 128-channel Hydrocel Geodesic Sensor Net (n = 845; ~94% of the current dataset). Study 2 used only the 128-channel net. All nets were connected to a NetAmps 200 or 300 amplifier (Study 1) or NetAmps 300 (Study 2) and sampled at 250 or 500 Hz. EEG was collected within an electrically shielded room.

#### NBS-Predict

To identify specific edge-level differences in groups a series of edge analysis were conducted using Network-Based Statistic-Predict (*NBS-Predict*; (Serin et al., 2021)). NBS-Predict is a prediction-based extension of the NBS framework designed to identify brain network connections that are predictive of behavioral or clinical outcomes. It integrates machine learning with cross-validation to directly assess the predictive utility of connectivity patterns at the network level, which improves model generalizability compared to the original NBS approach. In brief, this method first fits a predictive model (classification or regression) using edge-wise connectivity values as features, with possible covariates (e.g., sex, study). Then, for each cross-validation fold, edges are thresholded based on their association with the outcome variable, and connected components (subnetworks) are identified. Model performance is then evaluated using cross-validation metrics (e.g., classification accuracy), and permutation testing is employed to assess the statistical significance of predictive performance while controlling for multiple comparisons across the network.

### SI2. Statistical Analyses

#### *Generalized additive mixed models (GAMMs)*

Models were fit using mgcv package(version 1.9-1) and R (version 4.3.2). A separate GAMM was fit to predict each measure (e.g. functional connectivity, clustering coefficient). First, to determine whether to include an age-by-outcome interaction, two models were fit with the following forms:

Model 1 = EEG Measure ~ oGroup + s(age days, k = 4) + s(StudyID, bs = ‘re’) + s(Study, bs = ‘re’) + sex

Model 2 = EEG Measure ~ oGroup + s(age days, k = 4) + s(age days, by = oGroup, k = 4) + s(StudyID, bs = ‘re’) + s(Study, bs = ‘re’) + sex

oGroup represents outcome group stored as an ordered factor. Coding outcome as an ordered factor is necessary for the GAMMs model to produce a single significance value for the age-by-outcome interaction, which would allow direct comparison of outputs between groups. s(age_days, k = 4, bs = 'tp') is a smoothed age term. Study and StudyID are each included as random effects to account for repeated observations and clustering of observations within studies. Sex was included as a covariate. Model 2 (including the interaction term) was chosen if the difference was significant ($p < 0.05$). To correct for multiple comparisons, the false discovery rate (FDR) was controlled using the Benjamini and Hochberg method, which was applied for each model term across the four measure types to produce q-values (FDR-corrected p-values).

**SI3. Results**

**Local features: functional connectivity by ROI**

Using GAMMs, we identified four regional pairs that exhibited statistically significant developmental differences in COH between the Autism and LL-noAutism groups (Supplementary Figure 5). During infancy, infants later diagnosed with autism showed significantly higher COH than their LL-noAutism peers in central-to-posterior (84–140 days) and posterior-to-posterior connections (84–255 days) (Figure 5B and D). However, after age two, this pattern reversed: children with Autism demonstrated lower COH relative to LL-noAutism in frontal-to-frontal (948–1210 days; $F = 7.6$, $q < 0.03$), central-to-posterior (790–1210 days; $F = 13.18$, $q < 0.005$), temporal-to-posterior (584–1210 days; $F = 19.32$, $q < 0.01$), and posterior-to-posterior (687–1210 days; $F = 24.15$, $q < 0.01$) regions. GAMM results indicated that this group difference was primarily driven by a reduction in COH over time in the Autism group, rather than an increase in the LL-no Autism group.

*Trajectory of NBS identified clusters.* Next, sought to explore and characterize the trajectory of each significant subnetwork across age. For instance, for the early hyperconnectivity subnetwork identified at 3 months, we wanted to characterize how connectivity within those clusters evolved over time. Similarly, for the later hypoconnectivity subnetwork identified at 36 months, we wanted to assess potential patterns that may emerge over a developmental time scale. For this analysis, we quantified connectivity within NBS-defined subnetworks by averaging across all off-diagonal entries within the corresponding black mask areas for this analysis (main text Figure 3A,C).

First, the trajectory of early hyperconnectivity at 3 months exhibited a significant age-by-outcome interaction ($F = 17.68$, $q < 0.001$). Autistic infants showed elevated connectivity during early infancy that persisted until approximately 8 months, with connectivity increasing modestly with age. Second, the subnetwork representing early-hypoconnectivity at 3m revealed a diverging pattern between Autism and LL-noAutism group, where autistic infants exhibited hypoconnectivity up until 9 months. Autism group exhibited a gradual increase during the first year, peaking in the second year followed by a decline. In contrast, the LL-noAutism group showed less changes across age, with a slight decrease followed by increase in the second year. Third, the trajectory of later-hypoconnectivity at 24 and 36 months showed patterns consistent with global measures. The LL-noAutism group showed a gradual increase during the first year, peaking in the second year. By contrast, the Autism group showed a ‘growth-then-decline’ trend.

**Supplemental Table 1**. Participant EEG data included in analysis

| Age Group | Autism | | LL-noAutism | |
|---|---|---|---|---|
| | N | Age (mean (SD)) | N | Age (mean (SD)) |
| 3 | 24 | 108.62 (14.95) | 48 | 103.92 (10.85) |
| 6 | 19 | 191.42 (8.9) | 53 | 195.34 (10.86) |
| 9 | 25 | 283.72 (10.28) | 58 | 283.81 (10.43) |
| 12 | 46 | 382.93 (16.43) | 173 | 392.06 (22.46) |
| 18 | 39 | 563.41 (12.59) | 130 | 564.05 (11.49) |
| 24 | 41 | 749.46 (22.6) | 117 | 749.03 (16.08) |
| 36 | 35 | 1117.71 (22.88) | 105 | 1117.72 (19.66) |

**Supplemental Table 2**. EEG data quality metrics

| **EEG quality metrics**, Mean (SD) | | | |
|---|---|---|---|
| | Combined | LL-noAutism | Autism |
| Number of Segments | 317.96 (151.24) | 320.13 (145.78) | 311.22 (168.26) |
| Percent Good Channels | 91.79 (4.41) | 91.90 (4.37) | 91.45 (4.55) |

**Supplemental Table 3**. Alpha Connectivity Peak Frequency by age group

| Age Group | Alpha Connectivity Peak Frequency | |
|---|---|---|
| | Range | Median |
| 6 | 6.2–9.6 | 7.5 |
| 9 | 6.4–9.6 | 7.4 |
| 12 | 6.2–9.6 | 7.6 |
| 18 | 6.2–9.4 | 8.0 |
| 24 | 6.2–9.6 | 8.2 |
| 36 | 6.2–9.6 | 8.4 |

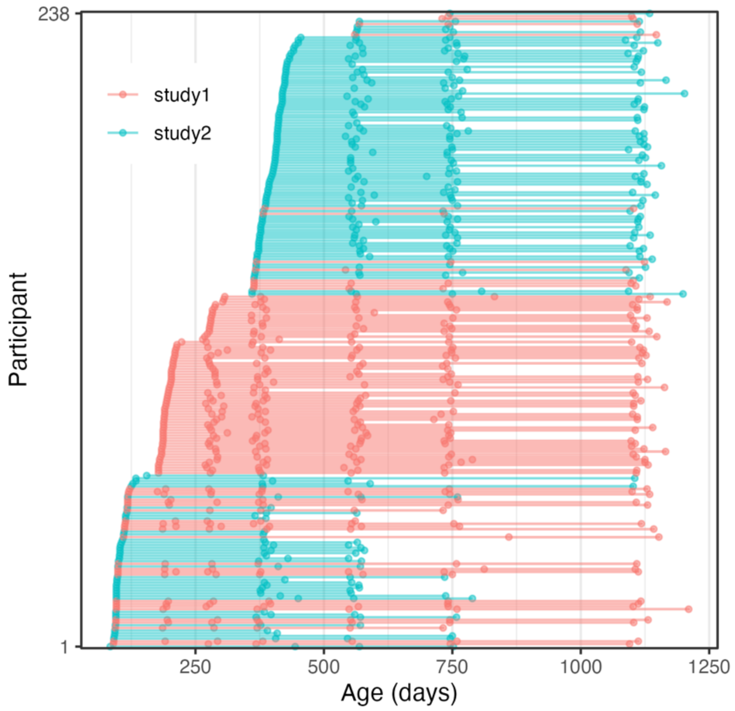


**Supplemental Figure 1.** Longitudinal study enrollment. Each line is a participant with dots indicating when EEG was collected for that participant.

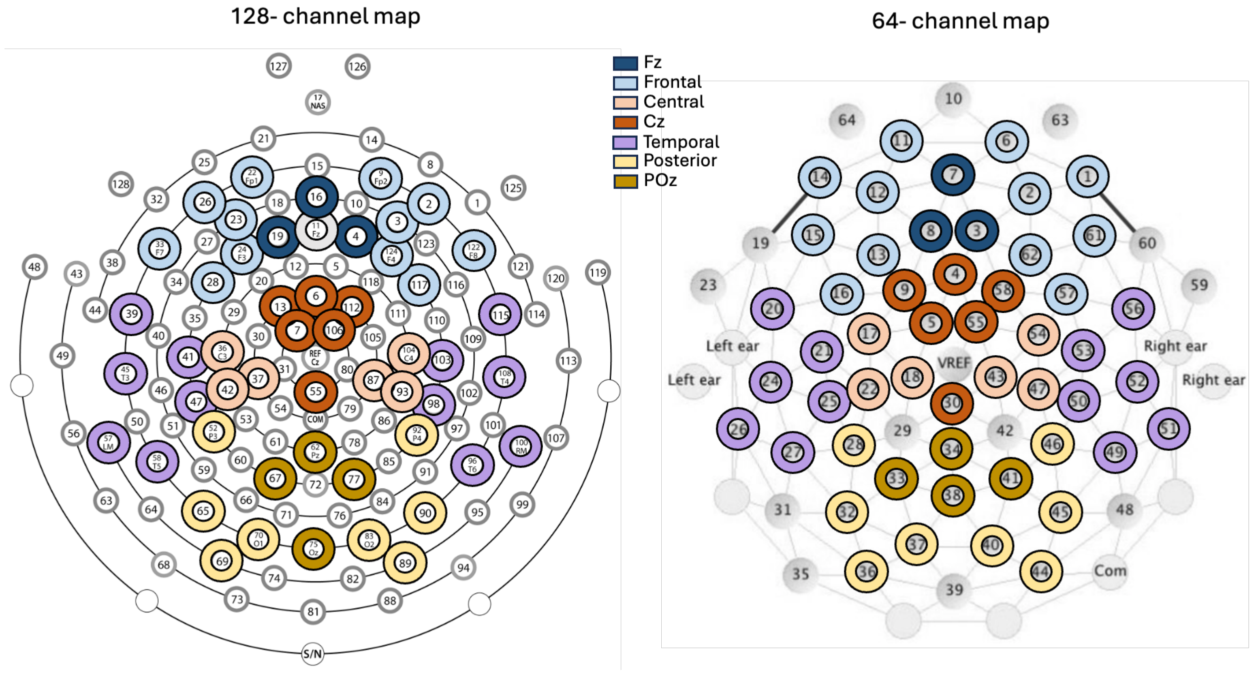


**Supplemental Figure 2.** Electrode layout: (A) 128-channel Hydrocel Geodesic Sensor Net. (B) 64-channel Geodesic Sensor Net. Electrodes averaged for Fz (navy blue), Frontal (light blue), Cz (dark orange), central (light orange), temporal (purple), and Pz (dark yellow), posterior-occipital (yellow) regions of interest.

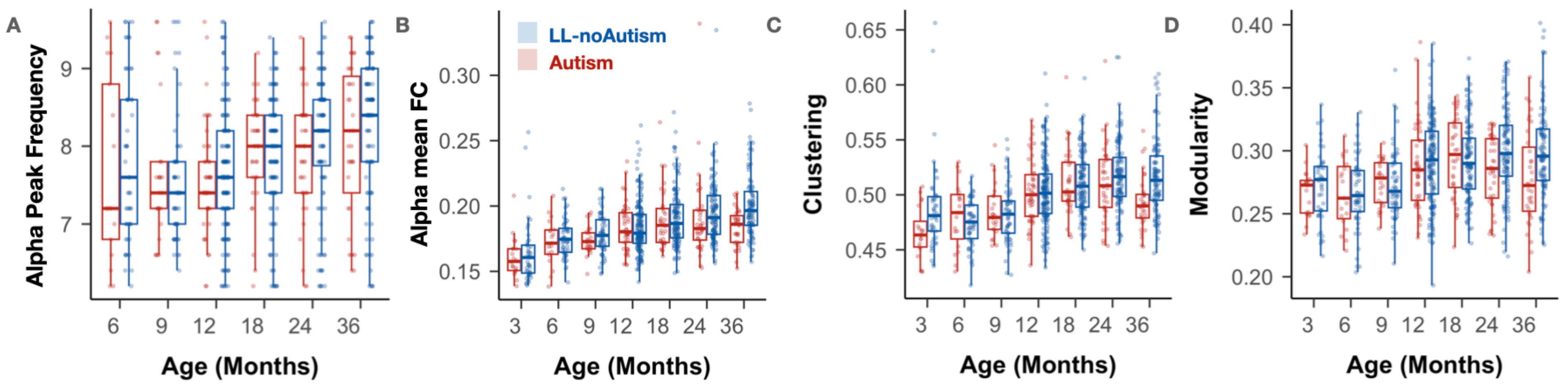


**Supplemental Figure 3:** Non-GAMMs modeled global measures of A) alpha peak frequency, B) alpha mean FC magnitude, C) clustering coefficient, and D) modularity.

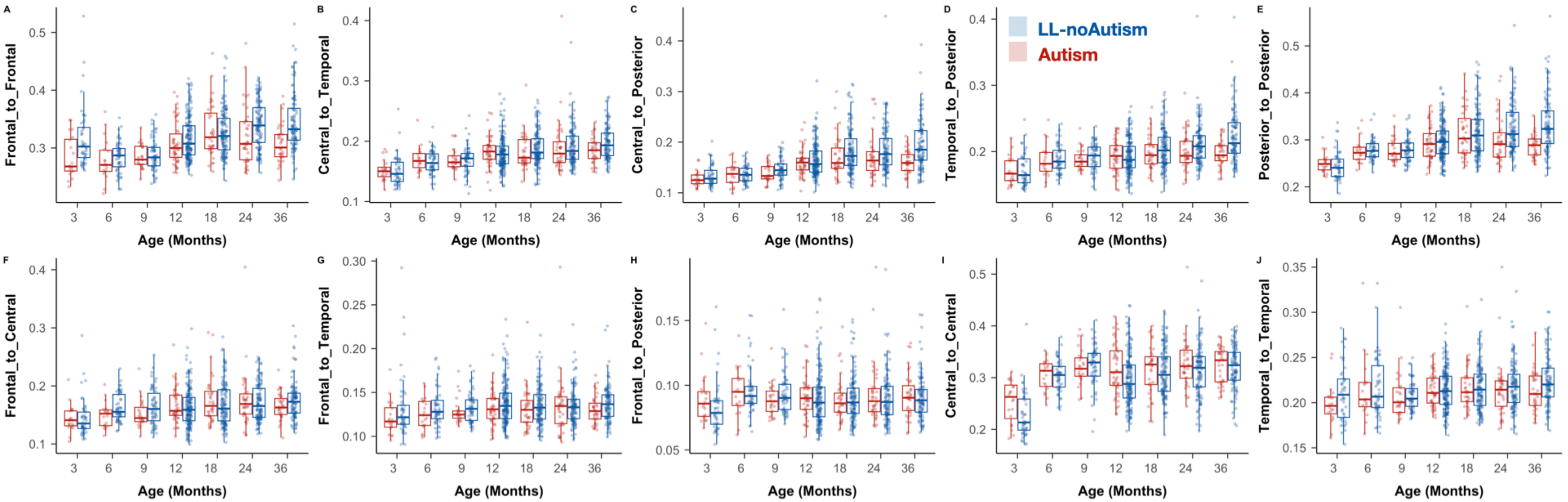


**Supplemental Figure 4:** Non-GAMMs local ROI-level measures of connectivity

**Supplementary Table 4**. Developmental trajectories of EEG measures and NBS-identified subnetworks by outcome: GAMM results

| FC features / NBS-Subnetworks | Age Effect F (q) | Age × Outcome Interaction F (q) | LL-noAutism Trajectory | Autism Trajectory | Group Divergence |
|---|---|---|---|---|---|
| **Global EEG Measures** | | | | | |
| **PACF** | F = 56.65, q < 0.001 | n.s. (p > 0.2) | Linear increase | Linear increase | None |
| **Mean Alpha FC** | F = 20.02, q < 0.001 | F = 7.84, q = 0.008 | Gradual increase | Increase to ~26 months, then decline (inflection: 807 days) | Autism ↓: 27 months (820 days) |
| **Clustering Coefficient** | F = 14.4, q < 0.001 | F = 4.11, q < 0.001 | Gradual increase; plateau 22-40 months (peak ~860 days) | Steeper early increase (peak: ~700 days); then decline | Autism ↑: 12-18 months (higher in Autism);<br>Autism ↓: 33-36 months |
| **Modularity** | F = 9.14, q < 0.001 | F = 7.2, q = 0.001 | Gradual increase | Steeper early increase (peak ~600days) then decline | Autism ↑: Up to 6.5 months<br>Autism ↓: 32-36 months (lower in Autism) |
| **NBS-Identified Subnetwork Trajectories** | | | | | |
| **Early hyperconnectivity (3m mask)** | F = 3.63, q = 0.057 | F = 17.68, q < 0.001 | Gradual increase | Elevated connectivity in early infancy; persisted until ~ 8 months (243 days) | Autism ↑: 3-8 months (~243 days) |
| **Early hypoconnectivity (3m mask)** | F = 12.42, q < 0.001 | F = 10.81, q < 0.001 | Inverted U-shape (decrease early until 13 months (393 days), then gradual increase) | Hyperconnectivity until 20 months (peak ~601 days); decline thereafter | Autism ↓: 3-9.2 months (~277 days) |
| **Later hypoconnectivity (24m mask)** | F = 2.52, q < 0.05 | F = 16.3, q < 0.001 | Gradual increase, peaking ~28 months (860 days) | Growth-then-decline: peak ~15 months (455 days) | Autism ↓: 19.5-36 months (585-1210 days) |
| **Later hypoconnectivity (36m mask)** | F = 9.95, q < 0.001 | F = 18.81, q < 0.001 | Gradual increase, peaking in year 2 | Growth-then-decline: peak ~ 23.3 months (700 days) | Autism ↓: 25.5.5-36 months (766-1210 days) |

*See main text Figures 4 and 5 for modeled trajectories.*

**Supplemental Table 5.** Nodes with significant differences and their degree from NBS-Predict (3mo)

A) Autism > LL-noAutism

| Region | Hemisphere | Electrode | Degree |
|---|---|---|---|
| frontal | left | E33 | 3 |
| frontal | left | E22 | 1 |
| frontal | left | E26 | 1 |
| frontal | right | E2 | 6 |
| frontal | right | E122 | 4 |
| frontal | right | E9 | 1 |
| central | left | E37 | 3 |
| central | left | E42 | 2 |
| central | left | E36 | 1 |
| central | right | E87 | 1 |
| temporal | left | E41 | 2 |
| temporal | left | E57 | 2 |
| temporal | left | E45 | 1 |
| temporal | left | E47 | 1 |
| temporal | left | E58 | 1 |
| temporal | right | E96 | 3 |
| posterior | left | E65 | 7 |
| posterior | left | E69 | 5 |
| posterior | left | E52 | 3 |
| posterior | middle | E62 | 6 |
| posterior | middle | E77 | 6 |
| posterior | middle | E67 | 5 |
| posterior | right | E90 | 5 |
| posterior | right | E83 | 2 |
| posterior | right | E89 | 2 |

Note: The weight threshold of 0.9 was applied.

B) LL-noAutism > Autism

| Region | Hemisphere | Electrode | Degree |
|---|---|---|---|
| frontal | left | E22 | 4 |
| frontal | left | E26 | 3 |
| frontal | left | E23 | 2 |
| frontal | left | E28 | 2 |
| frontal | left | E33 | 2 |
| frontal | left | E24 | 1 |
| frontal | middle | E16 | 3 |
| frontal | middle | E19 | 3 |
| frontal | middle | E4 | 1 |
| frontal | right | E122 | 6 |
| frontal | right | E2 | 3 |
| frontal | right | E3 | 3 |
| frontal | right | E117 | 2 |
| frontal | right | E124 | 1 |
| central | left | E42 | 2 |
| central | left | E37 | 1 |
| central | left | E36 | 1 |
| central | right | E93 | 2 |
| temporal | left | E39 | 7 |
| temporal | left | E45 | 5 |
| temporal | left | E58 | 3 |
| temporal | left | E57 | 2 |
| temporal | left | E41 | 1 |
| temporal | right | E108 | 7 |
| temporal | right | E96 | 5 |
| temporal | right | E115 | 3 |
| temporal | right | E100 | 2 |
| posterior | left | E52 | 5 |
| posterior | left | E69 | 3 |
| posterior | left | E65 | 1 |
| posterior | left | E70 | 1 |
| posterior | middle | E75 | 6 |
| posterior | middle | E62 | 2 |
| posterior | right | E83 | 2 |
| posterior | right | E89 | 1 |
| posterior | right | E92 | 1 |

**Supplemental Table 6.** Nodes with significant differences and their degree from NBS-Predict (24mo: LL-noAutism > Autism)

| Region | Hemisphere | Electrode | Degree |
|---|---|---|---|
| frontal | left | E23 | 5 |
| frontal | left | E26 | 3 |
| frontal | left | E24 | 2 |
| frontal | left | E33 | 2 |
| frontal | left | E22 | 1 |
| frontal | left | E28 | 1 |
| frontal | middle | E19 | 2 |
| frontal | middle | E4 | 1 |
| frontal | right | E2 | 4 |
| frontal | right | E3 | 4 |
| frontal | right | E9 | 4 |
| frontal | right | E124 | 1 |
| central | right | E93 | 4 |
| central | right | E87 | 2 |
| central | right | E104 | 1 |
| temporal | left | E45 | 4 |
| temporal | left | E58 | 2 |
| temporal | left | E57 | 1 |
| temporal | right | E100 | 4 |
| temporal | right | E98 | 3 |
| temporal | right | E103 | 2 |
| temporal | right | E115 | 2 |
| temporal | right | E108 | 2 |
| temporal | right | E96 | 1 |
| posterior | left | E65 | 3 |
| posterior | left | E70 | 3 |
| posterior | left | E69 | 2 |
| posterior | left | E52 | 1 |
| posterior | middle | E75 | 3 |
| posterior | middle | E77 | 2 |
| posterior | middle | E62 | 1 |
| posterior | middle | E67 | 1 |
| posterior | right | E89 | 8 |
| posterior | right | E83 | 6 |
| posterior | right | E90 | 6 |

Note: The weight threshold of 0.9 was applied.

**Supplemental Table 7**. Nodes with significant differences and their degree from NBS-Predict (36mo: LL-noAutism > Autism)

| Region | Hemisphere | Electrode | Degree |
|---|---|---|---|
| frontal | left | E22 | 11 |
| frontal | left | E23 | 6 |
| frontal | left | E26 | 5 |
| frontal | left | E24 | 5 |
| frontal | left | E28 | 2 |
| frontal | left | E33 | 1 |
| frontal | middle | E19 | 4 |
| frontal | middle | E4 | 3 |
| frontal | right | E2 | 6 |
| frontal | right | E3 | 6 |
| frontal | right | E124 | 4 |
| frontal | right | E9 | 2 |
| frontal | right | E122 | 2 |
| frontal | right | E117 | 2 |
| central | left | E37 | 1 |
| central | left | E36 | 1 |
| central | right | E87 | 12 |
| central | right | E93 | 9 |
| temporal | left | E58 | 8 |
| temporal | left | E41 | 6 |
| temporal | left | E45 | 5 |
| temporal | left | E57 | 5 |
| temporal | left | E39 | 2 |
| temporal | left | E47 | 2 |
| temporal | right | E108 | 7 |
| temporal | right | E100 | 4 |
| temporal | right | E103 | 2 |
| temporal | right | E96 | 2 |
| temporal | right | E98 | 1 |
| posterior | left | E69 | 9 |
| posterior | left | E70 | 9 |
| posterior | left | E52 | 7 |
| posterior | left | E65 | 7 |
| posterior | middle | E75 | 13 |
| posterior | middle | E62 | 9 |
| posterior | middle | E77 | 9 |
| posterior | middle | E67 | 8 |
| posterior | right | E83 | 7 |
| posterior | right | E89 | 6 |
| posterior | right | E92 | 6 |
| posterior | right | E90 | 4 |

Note: The weight threshold of 0.9 was applied.

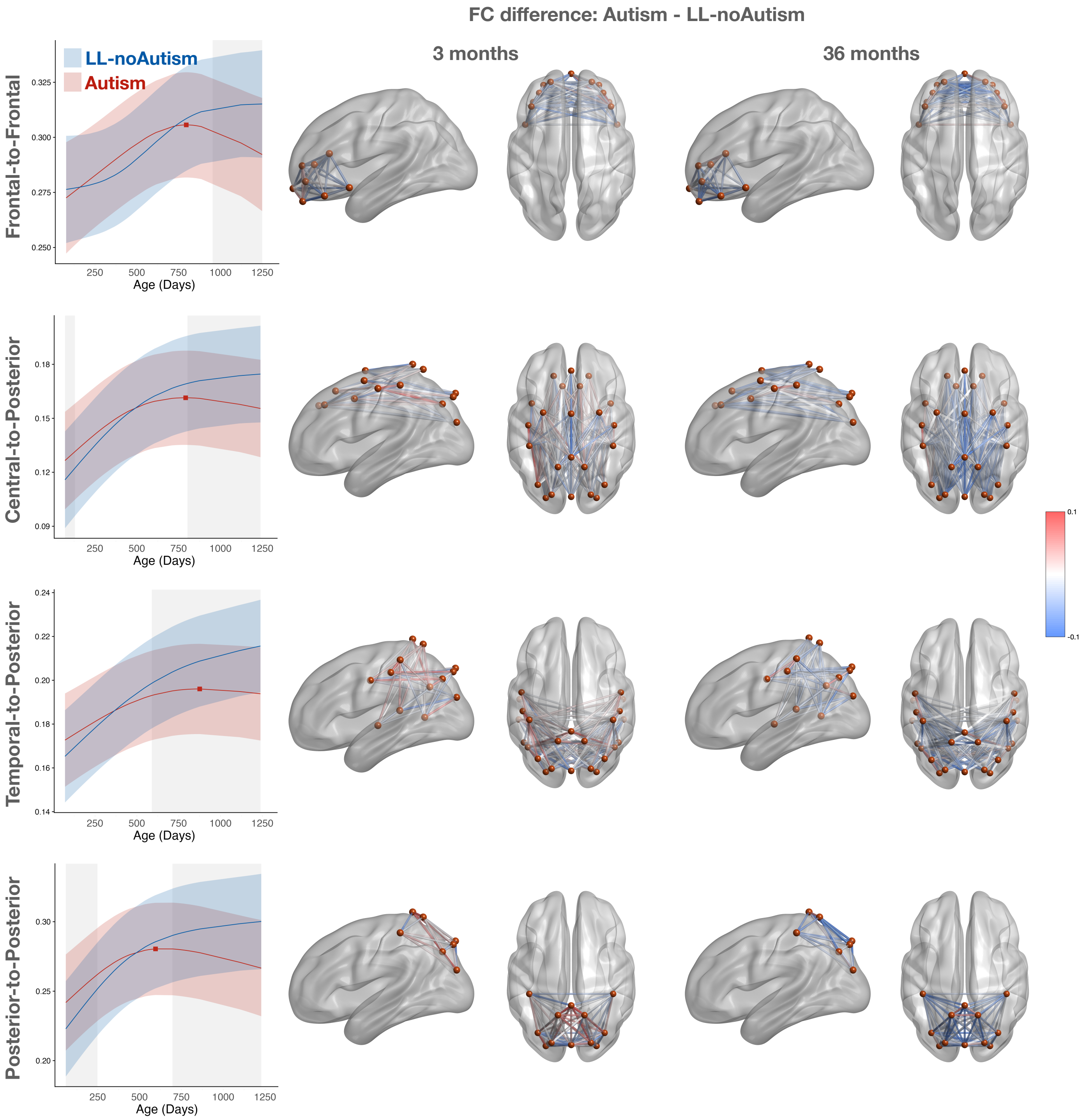


**Supplemental Figure 5.** GAMMs modeled trajectories for LL-noAutism (blue) and Autism (red) for connectivity in A) frontal-to-frontal, B) central-to-posterior, C) temporal-to-posterior, and D) posterior-to-posterior. COH values were averaged across all off-diagonal entries within the corresponding black mask areas for this analysis. Lines are the modeled mean predicted value with the shaded area representing 95% confidence intervals.
